# Assessing Lipid Core Burden Index with Depolarization-Sensitive Optical Frequency Domain Imaging

**DOI:** 10.64898/2026.05.22.26353889

**Authors:** Georgia L. Jones, Kenichiro Otsuka, Naoki Fujisawa, Hiroki Yamaura, Kotaro Matsumoto, Akihiro Okamoto, Tomohiro Yamaguchi, Takenobu Shimada, Shunsuke Kagawa, Takanori Yamazaki, Takashi Akasaka, Brett E. Bouma, Martin Villiger, Daiju Fukuda

**Author notes:** Authors contributed equally (Kenichiro Otsuka, Martin Villiger). **CORRESPONDING AUTHORS:** Martin Villiger Wellman Center for Photomedicine, Massachusetts General Hospital, Somerville, Massachusetts, 02145 **Email:**, Kenichiro Otsuka, Department of Cardiovascular Medicine, Osaka Metropolitan University Graduate, School of Medicine, Osaka, Japan, **Email:**.

## Abstract

**Background:** Quantitative lipid assessment is central to identifying rupture-prone coronary plaques and represents a therapeutic target for lipid-lowering therapy. Near-infrared spectroscopy (NIRS)-derived lipid core burden index (LCBI) is well validated and widely used for detecting lipid-rich lesions. Optical frequency domain imaging (OFDI) is increasingly adopted for guiding percutaneous coronary intervention (PCI) due to its high-resolution structural imaging capabilities. Depolarization-sensitive OFDI (depOFDI) provides intrinsic lipid contrast and may enable combined structural and compositional plaque characterization within a single OFDI-based platform.

**Objective:** To define an OFDI-derived lipid metric and evaluate its agreement with NIRS-derived LCBI.

**Methods:** Thirty-three patients underwent both polarization-sensitive OFDI and NIRS-intravascular ultrasound imaging during PCI. After exclusion of 4 datasets, 29 co-registered pullbacks were analyzed. A signal-to-noise-corrected depolarization metric was used to identify lipid-rich regions and generate depOFDI chemograms. maxLCBI_4mm_ value and location, as well as total LCBI, were computed and compared with NIRS.

**Results:** depOFDI demonstrated strong agreement with NIRS, showing high correlation for maxLCBI_4mm_ (r² = 0.862) and total LCBI (r² = 0.867), along with strong spatial concordance for the location of the maxLCBI_4mm_ (r² = 0.900). Bland-Altman analysis of LCBI_4mm_ showed minimal bias (10.7) with 95% limits of agreement of −81.4 to 102.8.

**Conclusions:** depOFDI enables accurate quantification of lipid burden alongside the high-resolution structural information inherently provided by OFDI. Because depolarization metrics can be derived from polarization-diverse detection available in many commercial OFDI systems, this approach provides a practical pathway toward comprehensive plaque characterization within existing PCI workflows, without the need for additional imaging modalities.

## INTRODUCTION

Atherosclerotic plaque rupture remains the predominant precipitating mechanism of acute coronary syndromes (ACS) [1,2]. While thin-cap fibroatheroma (TCFA) has historically served as a canonical morphological substrate of plaque vulnerability, contemporary intravascular imaging studies have demonstrated that no single structural phenotype fully captures rupture-prone biology [3–6]. Instead, quantitative imaging biomarkers, particularly those reflecting lipid burden, lipid composition, and microstructural collagen organization, are increasingly recognized as clinically relevant surrogates of plaque instability that can be measured in vivo [7–13].

Large prospective intracoronary imaging studies, including the PROSPECT and PREVENT trials, have established several quantitative predictors of future adverse cardiovascular events, such as minimal lumen area, plaque burden greater than 70 percent, OCT- and IVUS-defined TCFA, and lipid core burden index (LCBI) [7,10,12,14]. Among these, lipid burden assessed by near-infrared spectroscopy (NIRS) has emerged as one of the most robust and reproducible markers of vulnerable plaque biology [7,10,12,15]. NIRS-derived LCBI, particularly maximum LCBI over 4 mm (maxLCBI_4mm_), correlates strongly with histological lipid content and has been associated with both periprocedural myocardial infarction and future coronary events [7,10,12,15–17]. These findings have fueled interest in identifying non-culprit, lipid-rich, high-risk plaques at the time of the initial percutaneous coronary intervention (PCI), to enable prospective treatment of these lesions [12]. However, NIRS provides limited structural context and is most effective when combined with an intravascular imaging modality, requiring dedicated multimodal instrumentation and catheters.

Intravascular imaging-guided PCI has been shown to improve clinical outcomes [18–21]. Among available modalities, optical frequency domain imaging (OFDI) provides the highest-resolution structural characterization of coronary plaques [22–24]. Beyond purely morphological assessment, polarization-sensitive extensions of OFDI enable probing of tissue microstructure [25], including collagen organization via birefringence effects [26] and detection of lipid through depolarization [11]. While quantitative birefringence imaging typically requires modification of the imaging console by enabling polarization modulation in the sample arm [11,27], depolarization metrics can be obtained using polarization-diverse detection alone. Depolarization reflects polarization scrambling caused by multiple light scattering, which is prevalent in lipid-rich necrotic core material [28]. This capability is already available in several commercial OFDI instruments, including the Terumo Lunawave platform [27]. Depolarization therefore represents a practical and immediately translatable contrast mechanism for assessing lipid-rich plaque biology within an established intravascular imaging workflow. However, despite prior qualitative associations between depolarization and lipid, a quantitative and clinically validated assessment of lipid burden based on depolarization has not yet been established.

In the present study, we leverage depolarization-capable OFDI (depOFDI) to define a quantitative metric of depolarization as a surrogate of lipid burden and evaluate its relationship to NIRS-derived LCBI. Our objective is to assess the feasibility and accuracy of a strategy for identifying lipid-rich, rupture-prone plaques relevant to prospective risk stratification of non-culprit lesions during PCI, using a strategy compatible with existing OFDI instrumentation. Establishing concordance between depolarization-based OFDI metrics and NIRS-derived lipid burden would enable comprehensive structural and compositional plaque characterization using a single, widely available imaging platform, and may support the development of streamlined, imaging-guided preventive strategies in coronary artery disease.

## 1. METHODS

### 2.1 Study Population

This study was conducted at Osaka Metropolitan University Hospital (Osaka, Japan). It was approved by the Ethical Committee of Osaka Metropolitan University Graduate School of Medicine (approval number: 2022-035, UMIN000046910). Written informed consent was obtained from all patients before study enrollment. All procedures were conducted in accordance with the Declaration of Helsinki.

Between February 2023 and June 2025, a total of 33 patients undergoing PCI or follow-up assessment of lesions after prior coronary revascularization were enrolled in this study. The exclusion criteria were (1) age of patients <20 or >90 years at the time of consent; (2) patients with hemodynamic instability requiring mechanical support, such as a ventilator, intra-aortic balloon pumping, or extracorporeal membrane oxygenation; (3) patients with severe renal dysfunction (estimated glomerular filtration rate <30 mL/min/1.73m^2^) without undergoing maintenance dialysis; (4) individuals with poor life expectancy; and (5) pregnant patients.

Conventional OFDI was performed as part of routine clinical care. NIRS-IVUS was performed at the discretion of the treating clinicians to further assess plaque vulnerability. NIRS-IVUS was performed when clinically indicated and technically feasible, with exclusion of lesions exhibiting severe calcification or complex morphologies, including chronic total occlusion or marked vessel tortuosity. The target vessels were additionally imaged with a custom-built PS-OFDI console. For patients with ACS, NIRS-IVUS and depOFDI imaging of the culprit lesion was performed after restoration of normal coronary flow (TIMI grade 3), achieved using either small-diameter balloon dilatation (≤2mm) and/or aspiration thrombectomy. For patients with chronic coronary syndrome, physiological assessment was performed for all target lesions unless the stenosis was ≥ 90 % in the coronary angiogram. Additionally, non-invasive functional test results were considered to determine the target lesion. Written informed consent was obtained specifically for the prospective registry of intracoronary PS-OFDI imaging.

Demographic, clinical, angiographic, and procedural data were collected from electronic medical records and stored in a secure database. Imaging analyses were performed offline by experienced investigators blinded to all clinical and procedural information.

### 2.2 NIRS Image Acquisition

We employed the Makoto NIRS-IVUS system (TVC Imaging System MC-10; Infraredx, a Nipro Company, Burlington, MA, USA), featuring combined rotational NIRS and IVUS imaging at a pullback speed of 0.5 mm/s. Imaging was performed using a 3.2 French dual-modality catheter (Dualpro) equipped with a 50 MHz ultrasound transducer. This system generates a spatial lipid-probability map (chemogram) from which the LCBI is computed, providing a quantitative measure indicating the presence of a lipid-core plaque over any designated vessel length, together with an intrinsically co-registered IVUS pullback.

### 2.3 depOFDI Image Acquisition

Intracoronary depOFDI was performed using commercial intravascular imaging catheters (FastView™, Terumo Corp., Tokyo, Japan) connected to a custom PS-OFDI console. The console was derived from a wavelength-swept OFDI platform and modified at Massachusetts General Hospital for polarization-sensitive intravascular imaging, incorporating a Terumo motor drive unit, consistent with previously described systems [27,29]. The system operated at a center wavelength of 1310 nm with a 105-nm tuning range and a sweep rate of 50 kHz, yielding an axial resolution of approximately 9 µm assuming a tissue refractive index of 1.34. The system incorporated polarization-diverse detection and modulated the input polarization state between consecutive depth profiles. However, only a single input polarization state was used for depOFDI to emulate existing conventional OFDI systems. The intravascular catheter was pulled back at 10 to 20 mm/s at the operator’s discretion during continuous injection of non-ionic contrast at 3 to 5 mL/s. Images were acquired at 50 frames/s, with 1024 radial scans per frame. Conventional OFDI intensity images were displayed in the catheterization laboratory to provide real-time guidance regarding coronary anatomy.

### 2.4 NIRS-IVUS and depOFDI Image Registration

IVUS and OFDI pullbacks were longitudinally registered using coronary artery landmarks, including side branches and calcifications. Only arterial segments imaged by both modalities were included in the final analysis. Angular co-registration between IVUS and OFDI was performed by selecting three to four cross-sectional frames containing clearly recognizable anatomical landmarks visible in both modalities and computing the mean angular offset across these matched sections to define the final rotational alignment. As lipid core burden index metrics are cross section-based [7,10,12], angular alignment was not required for quantitative comparison.

### 2.5 depOFDI Chemogram Generation

Signal-to-noise ratio (SNR) and depolarization (DEP) images were generated from measurements corresponding to a single input polarization state using our previously described signal processing pipeline [27]. Restricting the analysis to a single input state is equivalent to operation with a conventional OFDI console. depOFDI processing was performed using spectral binning, in which the interferometric signal was divided into multiple spectral bins and the Stokes vector was calculated independently for each bin to improve polarization sensitivity [30].

For each bin, the complex-valued tomograms from the two polarization-diverse detection channels were converted into a four-component Stokes vector, **s** = [*I*, *Q*, *U*, *V*]^T^. This vector was then laterally filtered using a Gaussian window with a width of 6 depth scans, generating the averaged Stokes vector 〈**s***_p_*〉= [〈*I_p_*〉,〈*Q_p_*〉,〈*U_p_*〉,〈*V_p_*〉]^T^, where〈·〉 denotes spatial averaging and p denotes the spectral bin. The filtered intensity term, 〈*I*〉, represents the average backscattered intensity across both polarization channels and all spectral bins, while depolarization (DEP) was similarly averaged over all bins [30]. Signal-to-noise ratio was then calculated as SNR = 〈*I*〉/*N*, where *N* represents the system noise floor, estimated globally for the imaging system and applied uniformly across all datasets. DEP is described as the complement to one of the Euclidean norm of the second through fourth components over the total intensity and describes the degree of polarization of the light [31], with 0 indicating fully polarized light and 1 describing fully unpolarized light:

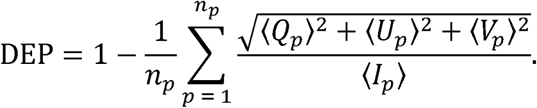

Where *n_p_* denotes the number of spectral bins. Theoretically, in the absence of intrinsic tissue depolarization, the SNR and DEP signals exhibit a predictable, theoretical relationship, as reductions in SNR lead to increases in depolarization solely due to measurement noise. This relationship can be described by the function

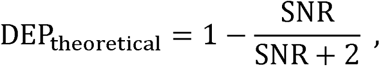

derived from system noise statistics [32,33]. A DEP value significantly lower than this bound indicates high likelihood for the presence of true depolarization attributable to microstructural or compositional heterogeneity within the tissue, such as lipid pools or necrotic cores.

To reliably judge whether the DEP value of a given voxel, together with its corresponding SNR, should count as depolarizing, we simulated the expected probability density function of the SNR-DEP relation by modeling fully polarized, fully developed speckle with varying mean signal amplitude and the addition of complex Gaussian noise in both polarization channels, and propagating these fields through Stokes parameter estimation and spatial averaging. Only DEP values with a probability <1% of being fully polarized were classified as exhibiting true depolarization, while those with a higher probability were considered non-depolarizing (as shown in Figure 1). Any points with an SNR value below 5 dB were determined to be noise dominated.

**Figure 1:**
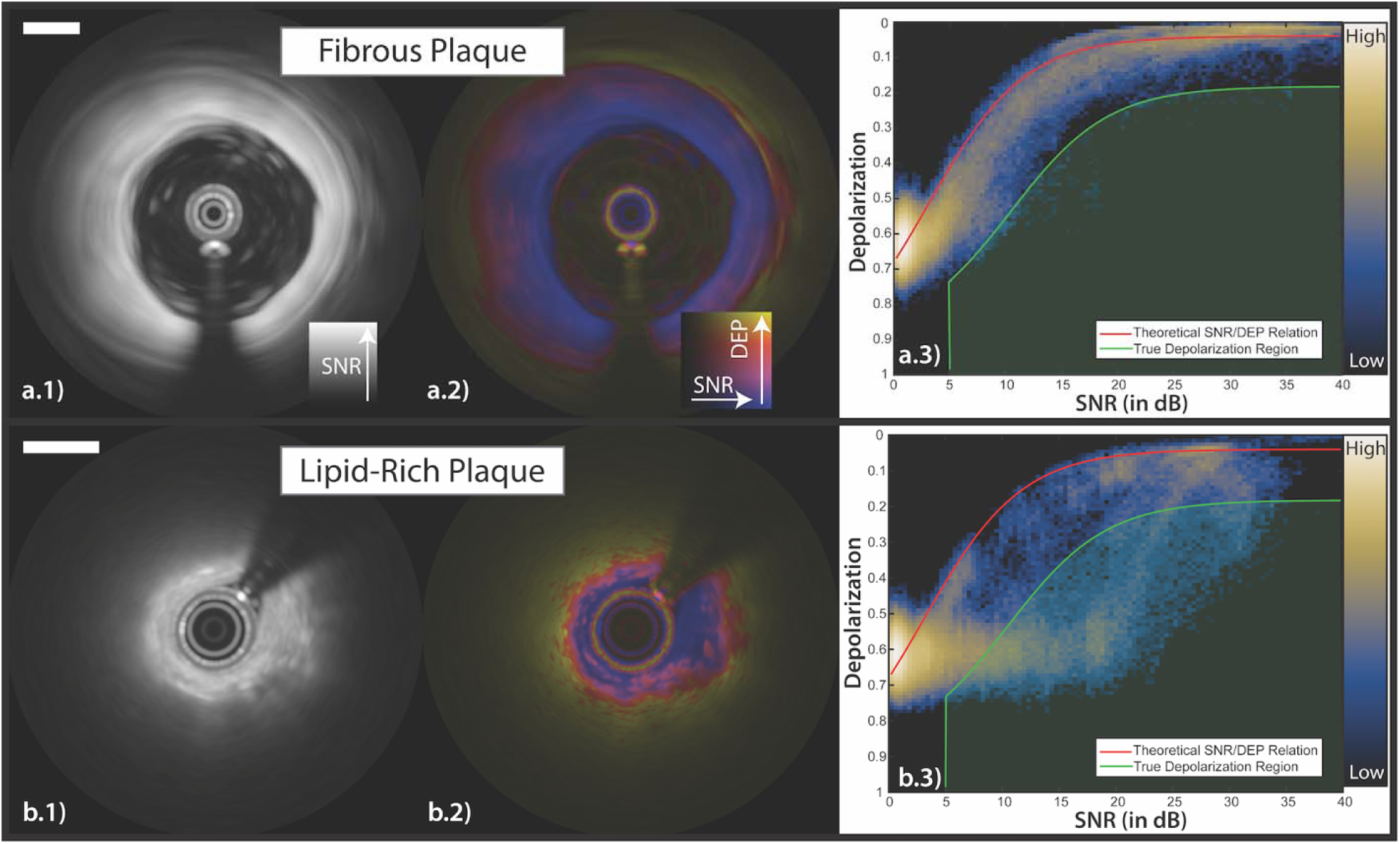
Representative example of lipid metric computation for regions without lipid (top row, a) and with lipid-rich plaque (bottom row, b). Corresponding filtered OFDI signal to noise ratio (SNR) images (a.1, b.1) and depolarization maps as color hue, overlayed with SNR images in brightness (a.2, b.2). Two-dimensional histograms (a.3, b.3) illustrating the relationship between SNR and DEP for all pixels in each frame. The theoretical SNR-DEP relationship (red curve) represents expected depolarization behavior solely due to noise in the absence of sample-induced depolarization, while the green area indicates pixels that are deemed “truly depolarized”. In the lipid-rich cross section (b), numerous points deviate substantially from the theoretical curve, indicating true depolarization consistent with lipid content. In contrast, the cross section with minimal lipid (a) closely follows the theoretical model, demonstrating minimal sample-induced depolarization. Scale bars: 1 mm.

Lipid detection was performed by analyzing spatial distributions of SNR-corrected DEP within a 10-depth-scan moving window (corresponding to an 8° rotation of the catheter). Within each window, the proportion of pixels exhibiting SNR-corrected depolarization was calculated as *N_dep_*⁄*N_tot_*, where *N_dep_* denotes the number of pixels below the SNR-DEP deviation threshold and *N_tot_* represents the total number of data points above the 5 dB SNR threshold in the window.

The resulting fraction of lipid containing pixels was expressed as a percentage and mapped along the vessel to generate a depOFDI-derived lipid chemogram, analogous to the chemograms produced by NIRS. The resulting chemograms were visualized using a color-coded overlay representing relative lipid burden. Points in the guidewire shadow were explicitly masked. Guidewire shadows were segmented using a laboratory-developed machine learning-based segmentation pipeline (OCTSeg) [34]. While OCTSeg requires polarization metrics, several published protocols exist for guidewire segmentation using OFDI intensity images alone [35–37]. The polarization metrics were computed in parallel to the depOFDI analysis for use in OCTSeg, as previously described [29,30]. Representative examples of depOFDI-derived chemograms and their corresponding NIRS-LCBI maps are shown in Figures 2 and 3.

**Figure 2:**
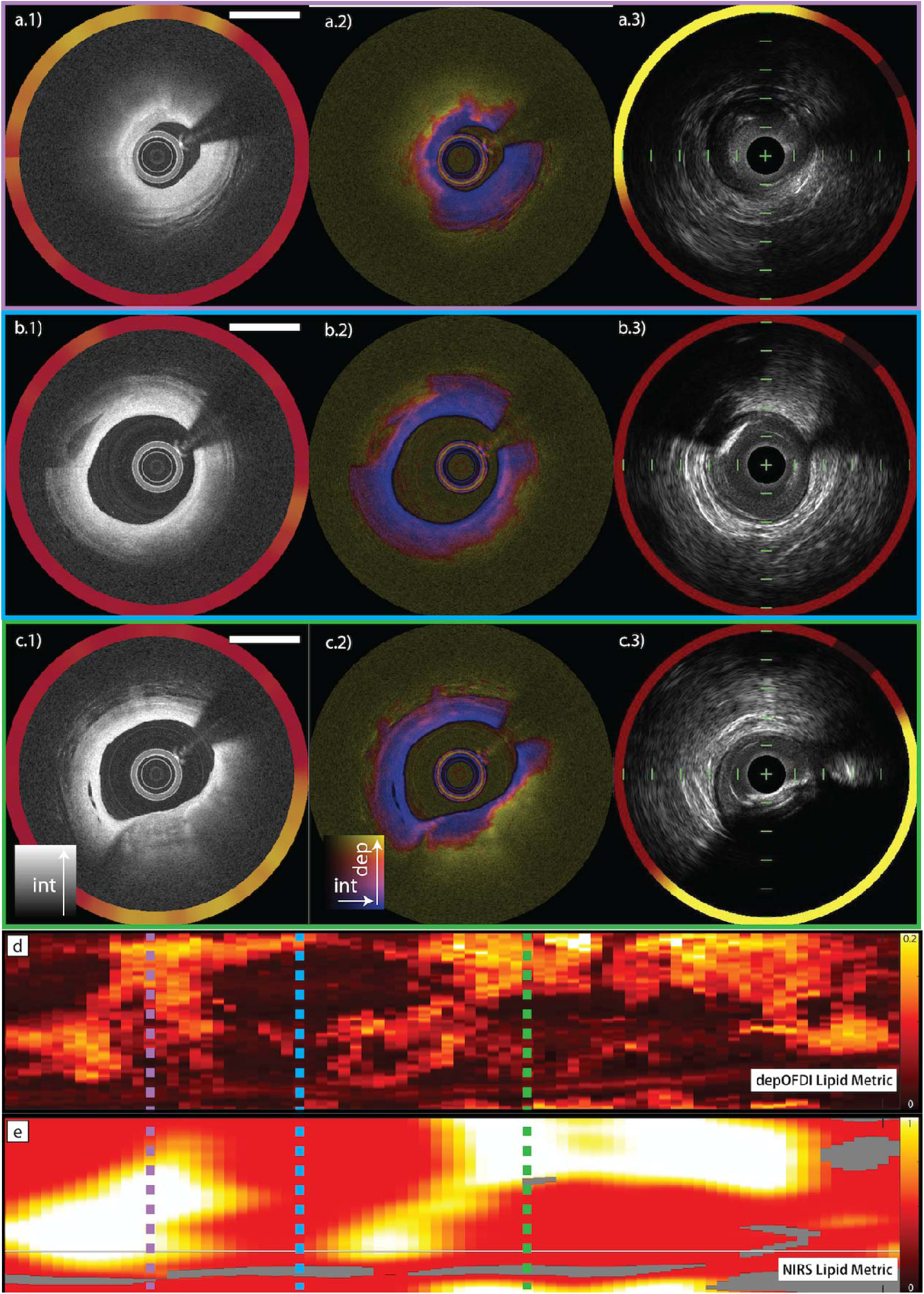
Representative example of matched near-infrared spectroscopy (NIRS) and depolarization-sensitive optical frequency domain imaging (depOFDI) pullbacks, highlighting three cross sections at distinct locations. depOFDI results are summarized as (a.1–c.1) OFDI intensity images with depOFDI-derived lipid metric ring and (a.2–c.2) corresponding depolarization images. The registered NIRS-IVUS cross sections are surrounded by the corresponding NIRS lipid probability (a.3–c.3). The longitudinal depOFDI-derived lipid metric chemogram generated from depolarization analysis (d) and the registered NIRS chemogram (e) illustrate strong spatial concordance between lipid-rich regions identified by either modality. Vertical dashed lines denote corresponding cross-sectional locations shown in panels (a–c). Scale bars (a-c): 1 mm.

**Figure 3:**
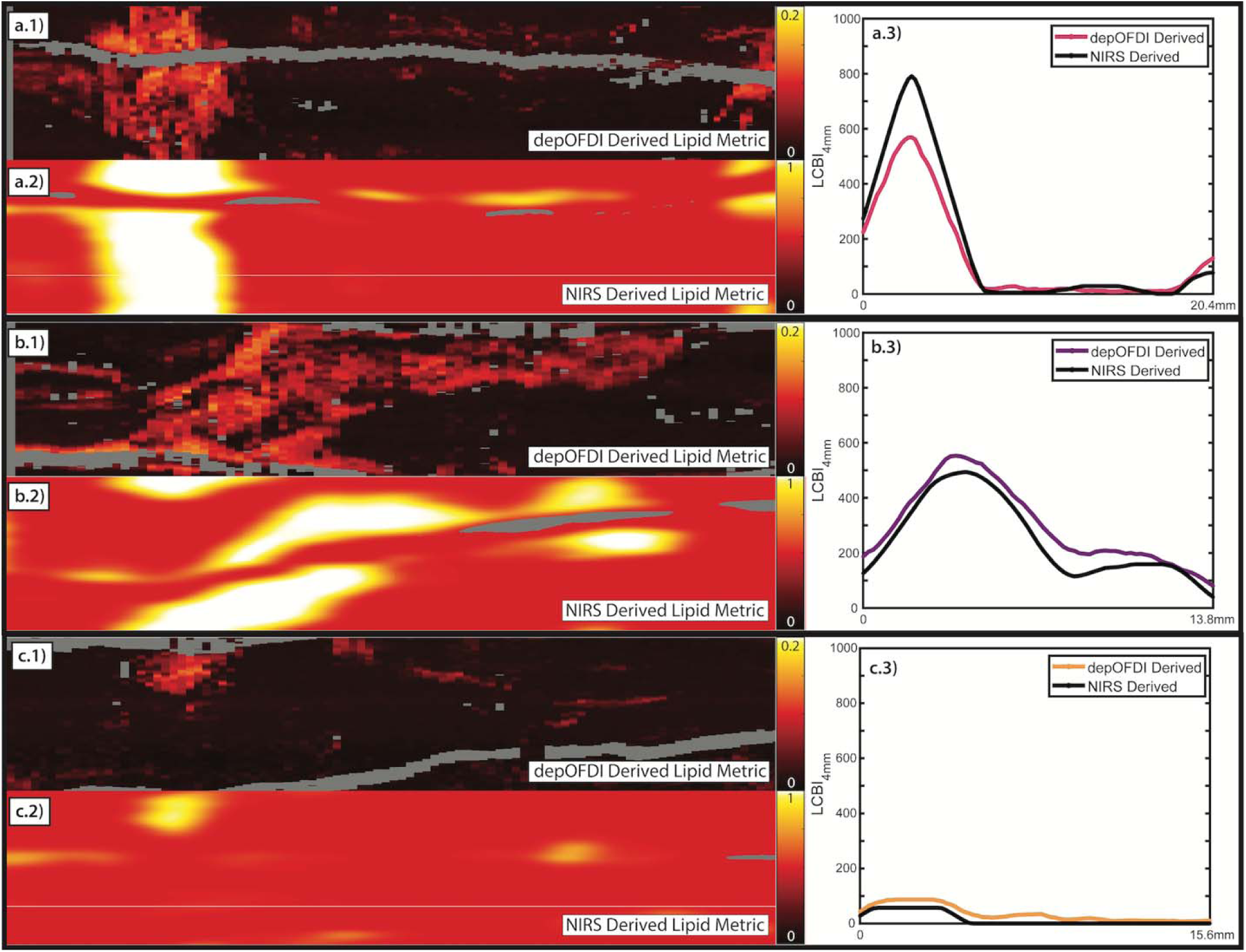
Comparisons between depOFDI-derived and NIRS-derived lipid chemograms for three representative coronary artery pullbacks. depOFDI-derived lipid metric chemograms generated using the SNR-corrected depolarization model (a.1–c.1) agree well with th corresponding NIRS-derived chemograms from the matching pullback segment (a.2–c.2). (a.3–c.3) Lipid core burden index computed over 4 mm segments (LCBI _mm_) for both depOFDI-derived (colored traces) and NIRS-derived (black traces) along the available pullback length.

### 2.6 Lipid Core Burden Index Computation

The LCBI was computed to quantify the extent of lipid-rich plaque along the imaged coronary segments. LCBI represents the percentage of lipid-containing area within a given pullback length in the chemogram, providing a continuous measure of lipid burden, as established by NIRS [7,10,12].

For NIRS imaging, the LCBI metrics were computed from the exported chemograms from the Infraredx system, which ranged from 0 to 1. The lipid threshold was calibrated by matching the totalLCBI computed from the exported chemogram to the totalLCBI value output in the system software. Using this method, the NIRS lipid threshold was determined to be 0.55. For the depOFDI-derived chemograms, lipid-containing regions were determined from the computed depolarization-based lipid metric. Each depOFDI-derived chemogram pixel was classified as lipid-containing if its lipid pixel ratio exceeded a threshold of >5%, which was determined using the mean of the minimum intra-class variance threshold as determined by Otsu’s method across all depOFDI chemograms. For both NIRS and depOFDI, pixels meeting the lipid criterion for the respective method were assigned a binary lipid value of one, and those below the threshold were assigned zero. The LCBI metrics were then calculated as the fraction of lipid-containing pixels within the specified pullback segment, expressed as a percentage of the total number of valid pixels multiplied by 1000, independently for both methods.

Furthermore, a local lipid core burden index over a 4-mm length (LCBI_4mm_) was computed using a sliding window along the pullback direction, based on the chemograms of either method. The window was translated in 0.2 mm increments across the entire imaged length, and the corresponding LCBI_4mm_ values were recorded. The maximum LCBI_4mm_ (maxLCBI_4mm_) was defined as the highest LCBI_4mm_ value observed across the matching pullback segments and used as the primary quantitative descriptor of local lipid burden. The maxLCBI_4mm_ location was defined as the longitudinal distance (in millimeters) from the proximal end of the co-registered segment corresponding to the center position of the window in which the maximum value occurred. The total LCBI was determined as the LCBI value when computed over the entire matching pullback segments.

### 2.7 Statistical Analysis

All statistical analyses were performed using MATLAB R2024b (MathWorks, Natick, MA, USA). Correlation between NIRS-derived and depOFDI-derived lipid metrics was evaluated on a per-patient basis using Spearman’s correlation coefficient. The squared correlation coefficient (r²) was computed for total LCBI, maxLCBI_4mm_, and the longitudinal position of the maxLCBI_4mm_ to assess agreement in lipid burden magnitude and spatial localization between the two imaging modalities. A p-value of <0.05 was considered to indicate statistical significance.

To further evaluate measurement agreement across the entire pullback, all available LCBI_4mm_ values were compared between depOFDI and NIRS using Bland-Altman analysis. The mean difference (bias) and 95% limits of agreement (mean difference ± 1.96 × standard deviation) were calculated to quantify systematic and random discrepancies between modalities.

## 2. RESULTS

### 3.1 Patient Characteristics

A total of 33 matching NIRS and depOFDI pullbacks were obtained from 33 patients who underwent clinically indicated PCI at Osaka Metropolitan University Hospital. Patient characteristics are described in Table 1. The mean age of the study population was 70.1 ± 10.4 years, and 75.8% were male. Four pullbacks were excluded due to poor OFDI image quality related to inadequate blood clearance and signal attenuation, resulting in a final dataset of 29 matched coronary vessel image data sets suitable for analysis.

**Table 1.** Patient characteristics.

| **Table 1. Patient characteristics** |  |
| --- | --- |
|  | N = 33 |
| Age (years), mean ± SD | 70.1 ± 10.4 |
| Male, n (%) | 25 (75.8%) |
| Body mass index, mean ± SD | 24.5 ± 3.5 |
| Diabetes, n (%) | 13 (39.4%) |
| Hypertension, n (%) | 27 (81.8%) |
| Dyslipidemia, n (%) | 29 (87.9%) |
| Smoking, n (%) | 15 (45.4%) |
| *Clinical presentation* |  |
| ACS n (%) | 10 (30.3%) |
| *Imaged vessel* |  |
| LAD, n (%) | 21 (63.6%) |
| RCA, n (%) | 8 (24.2%) |
| LCX, n (%) | 3 (9.1%) |
| *Laboratory test* |  |
| LDL cholesterol (mg/dL), mean ± SD | 100.1 ± 35.0 |
| HDL cholesterol (mg/dL), mean ± SD | 48.7 ± 11.0 |
| Triglyceride (mg/dL), mean ± SD | 153.9 ± 137.3 |
| Hemoglobin A1c (%), mean ± SD | 6.14 ± 0.6 |
| *Medical history* |  |
| Chronic kidney disease, n (%) | 12 (36.4%) |
| Prior myocardial infarction, n (%) | 6 (18.2%) |
| Prior PCI, n (%) | 6 (18.2%) |
| Prior CABG, n (%) | 1 (3.0%) |
| Abbreviations: ACS, acute coronary syndrome; PCI, percutaneous coronary intervention; CABG, coronary artery bypass grafting. |  |

The majority of lesions were located in the left anterior descending artery (n = 21, 63.6%) and 30.3% of cases presented with ACS, while 69.7% were classified as chronic coronary syndrome. All included cases demonstrated adequate overlap between NIRS-IVUS and OFDI imaging pullbacks to permit precise co-registration and quantitative comparison.

### 3.2 Co-Registration and Qualitative Chemogram Comparison

The IVUS and OFDI volumes were longitudinally and rotationally co-registered to identify matching pullback segments. After registration, depOFDI-derived chemograms were generated using the lipid fraction metric, as described in Figure 1. Across all analyzed pullbacks, depOFDI-derived chemograms demonstrated strong qualitative agreement with NIRS chemograms. Regions of elevated depolarization-based lipid metric from depOFDI consistently matched with areas of high lipid probability on NIRS. Figure 2 presents a representative co-registered pullback, illustrating spatial alignment of lipid-rich regions between modalities. Matched cross-sectional images at selected chemogram locations demonstrate concordance between the depOFDI images and the corresponding registered NIRS-IVUS image both in vessel morphology and lipid pool location. Figure 3 displays LCBI_4mm_ values from three representative, co-registered depOFDI and NIRS pullbacks, demonstrating qualitative agreement in the longitudinal pattern and magnitude of lipid burden across the imaged segments.

### 3.3 Lipid Core Burden Index Comparison

Quantitative comparison demonstrated strong correlation between depOFDI-derived and NIRS-derived measures of lipid burden (Figure 4). Total LCBI and maxLCBI_4mm_ values exhibited significant Spearman correlation across the study population (r² = 0.862, p = 1.9•10^-9^ for maxLCBI_4mm_; r² = 0.867, p = 1.1•10^-9^ for total LCBI). The maxLCBI_4mm_ position along the pullback also showed good spatial agreement between modalities (r² = 0.900, p = 2.6•10^-11^), indicating consistent localization of the most lipid-rich plaque regions.

**Figure 4:**
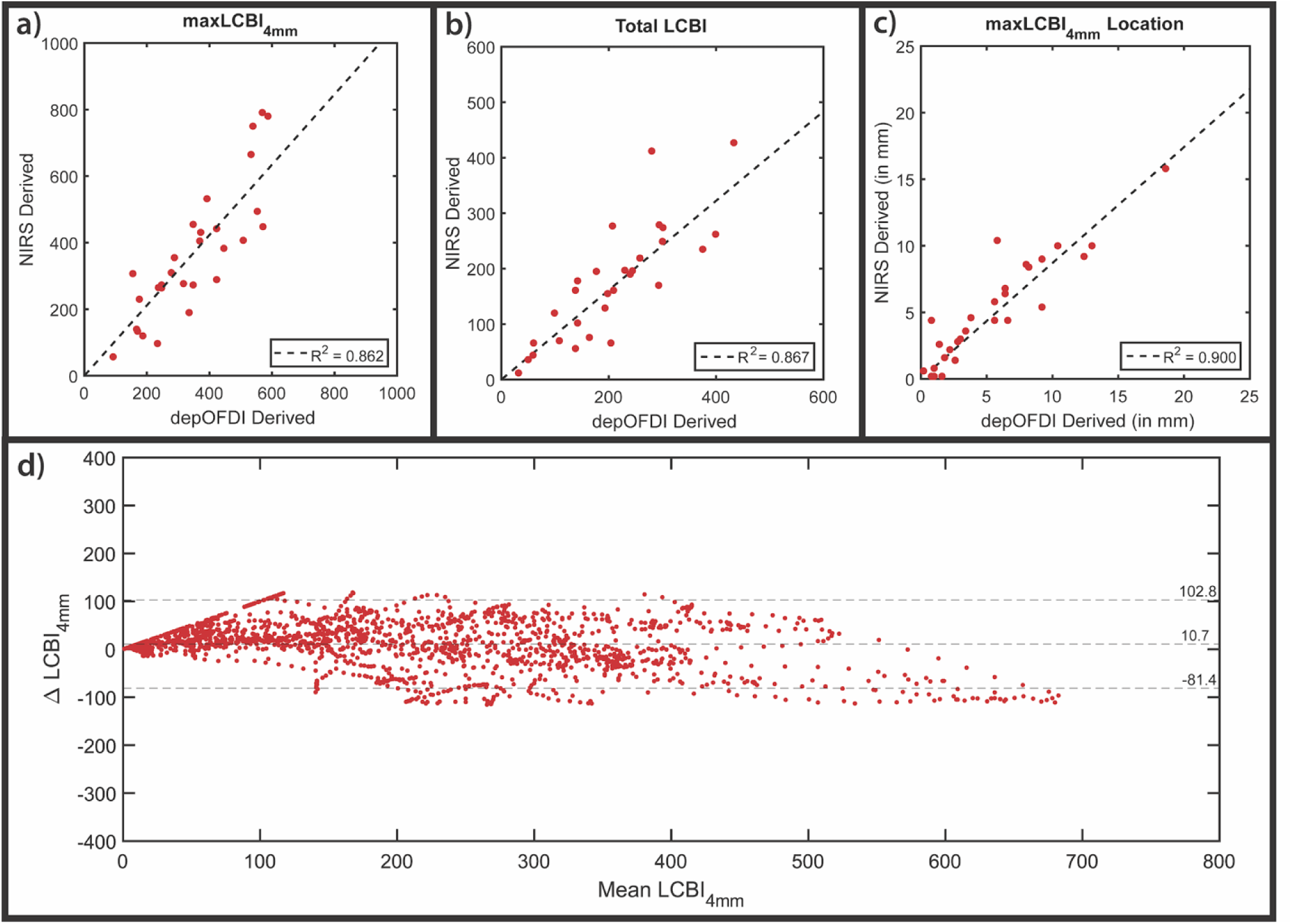
Comparison of depOFDI-derived and NIRS-derived lipid core burden measurements across all matched pullback segments. (a–c) Scatterplots of (a) maxLCBI _mm_, (b) total LCBI, and (c) maxLCBI _mm_ location for all patients. (d) Bland-Altman plot, illustrating the mean bia (10.7) and 95% limits of agreement ([−81.4,102.8]) of all recorded LCBI_4mm_ measurements (n = 2189) between the two modalities. Each data point represents an LCBI_4mm_ measurement, which is computed with a sliding window across the entire matched pullbacks.

Bland-Altman analysis between all depOFDI-derived and NIRS-derived LCBI_4mm_ (n = 2189) values revealed minimal bias in the LCBI_4mm_ of 10.7, with depOFDI overestimating the LCBI relative to NIRS. The 95% limits of agreement ranged from −81.4 to 102.8 indicating good overall agreement. Graphical analyses and plots, including correlation scatterplots and Bland-Altman plot, are presented in Figure 4.

## 3. DISCUSSION

Lipid detection and plaque characterization have emerged as areas of increasing clinical importance, driven by the recognition that large lipid-rich atherosclerotic plaques underlie many acute coronary events [3–6]. Quantitative assessment of lipid content has been shown to provide valuable clinical information, including prediction of future adverse cardiovascular events, identification of lesions at risk for slow flow during PCI, and guidance for selecting optimal stent landing zones [7,10,12,15–17,38,39]. Over the past decade, NIRS has played a pivotal role in this domain, demonstrating strong predictive value for future adverse cardiovascular events through quantification of lipid content using the LCBI [7,10,12]. However, despite its demonstrated utility, the clinical adoption of NIRS-IVUS has remained limited. In addition to the need for a dedicated imaging catheter and proprietary console, the NIRS-IVUS catheter has a relatively large profile and suboptimal crossing performance, which can make its use challenging in tortuous vessels, severe stenoses, or calcified lesions. In contrast, intravascular imaging to guide PCI with smaller-profile IVUS and OFDI/OCT catheters is increasingly integrated into clinical practice.

In the present study, we demonstrated that depOFDI enables identification of lipid-rich regions within coronary arteries, with strong agreement with NIRS in both qualitative assessment and quantitative measures, including total LCBI and maxLCBI_4mm_. The spatial localization of lipid-rich segments also showed close correspondence between modalities. These findings indicate that depOFDI can provide a reliable surrogate for NIRS in quantifying lipid burden. By enabling simultaneous assessment of plaque structure, cap thickness, macrophage accumulation, and lipid content, depOFDI has the potential to deliver comprehensive coronary plaque characterization using a single imaging platform. This could substantially simplify procedural workflows, reduce cost, and facilitate broader adoption of advanced plaque imaging in routine clinical care.

A major practical advantage of depOFDI is its compatibility with existing commercial OFDI systems currently deployed in catheterization laboratories. The depOFDI framework requires no console modifications, specialized catheters, or hardware upgrades, leveraging polarization-diverse detection already present in many systems, such as the Terumo Lunawave [27]. While we used a custom PS-OFDI console for this initial study, analysis was restricted to a single input polarization state, rendering it conceptually equivalent to a conventional OFDI system. depOFDI may therefore be applied retrospectively on exported complex-valued tomogram data from polarization-diverse detection channels. Alternatively, the relatively modest computational requirements of depolarization metrics allow integration into standard reconstruction pipelines, facilitating translation into clinical research and practice.

Because NIRS detects diffusely scattered light, its detection sensitivity likely extends to larger penetration depths than the confocal OFDI signal. NIRS may therefore detect lipid pools located deeper within the vessel wall. However, lipid located closer to the luminal surface is likely of greater clinical relevance due to higher rupture risk. Accordingly, OFDI-derived depolarization may preferentially highlight superficial lipid-rich components more directly linked to adverse coronary events, whereas NIRS captures deeper lipid burden that may be less immediately associated with plaque instability.

Conventional OFDI demonstrates high sensitivity for detecting lipid-rich plaques, but its clinical utility is limited by a substantial false positive rate that reduces positive predictive value [28,40,41]. Many of these false positives arise from macrophage accumulation, as superficial macrophage accumulations produce signal attenuation patterns that are indistinguishable from necrotic lipid cores using intensity-based OFDI alone [28,40]. depOFDI may help overcome this limitation by detecting the pronounced depolarization of light caused by multiple scattering within lipid-rich necrotic core material, whereas superficial foamy macrophages exhibit only a modest depolarization signature.

This study should be interpreted considering several limitations. First, the sample size was small, and all patients were enrolled from a single center, which may limit the generalizability of the findings. Larger, multi-center studies are warranted to confirm the reproducibility of the observed correlations between depOFDI- and NIRS-derived lipid metrics. Second, although NIRS was used as the reference standard for lipid quantification, it represents an indirect measure of lipid content. Histopathologic validation of depOFDI would strengthen our findings. Finally, co-registration between NIRS and OFDI pullbacks was based on anatomical landmarks rather than precise fiducial markers, which introduces potential for small longitudinal misalignments in region-based comparisons. Future studies with larger, multi-center cohorts are warranted to validate these findings across diverse patient populations and imaging systems, and to evaluate the prognostic value of depOFDI-derived lipid indices for predicting clinical outcomes.

In conclusion, this study demonstrates that depolarization-sensitive OFDI enables robust quantification of coronary lipid burden with strong agreement to established NIRS-derived LCBI metrics. By leveraging polarization-diverse detection already available in existing OFDI systems, the proposed depOFDI framework provides the foundation for a practical, hardware-compatible approach for lipid characterization without the need for additional instrumentation.

### Clinical Perspectives

#### Clinical Competencies

This study establishes depOFDI as a quantitative, readily available method for lipid burden assessment with strong concordance to NIRS-derived LCBI. This may enable interventionalists to perform comprehensive plaque characterization within routine clinical workflows without additional hardware or specialized catheters.

#### Translational Outlook

Although this single-center study demonstrates strong technical agreement with NIRS, several key barriers must be addressed for clinical translation: (1) integration of depOFDI analysis into commercial OFDI consoles with real-time processing capability; and (2) histopathologic correlation studies to validate the depolarization-lipid relationship.

## Data Availability

All data produced in the present study are available upon reasonable request to the authors

## Acknowledgements

The authors used AI-assisted language editing for drafting assistance. All scientific content, data analysis, and conclusions were developed and verified by the authors.

## Nonstandard Abbreviations and Acronyms

ACS: acute coronary syndrome
CCS: chronic coronary syndrome
DEP: depolarization
depOFDI: depolarization-sensitive optical frequency domain imaging
IVUS: intravascular ultrasound
LCBI: lipid core burden index
NIRS: near-infrared spectroscopy
OFDI: optical frequency domain imaging
PS-OFDI: polarization-sensitive OFDI
TCFA: Thin-Cap Fibroatheroma

